# CYP2D6 variants in amyotrophic lateral sclerosis: an association study of risk and survival

**DOI:** 10.1101/2025.09.29.25336320

**Authors:** Johanna K Vallikivi, Maarten Kooyman, Project MinE ALS Sequencing Consortium, Alfredo Iacoangeli, Ammar Al-Chalabi, Ahmad Al Khleifat

## Abstract

Amyotrophic lateral sclerosis (ALS) is a progressive neurodegenerative disorder with limited therapeutic options. Riluzole remains the only widely available treatment in ALS, yet its benefits are modest and highly variable across patients. Genetic variation in cytochrome P450 2D6 (CYP2D6), a major enzyme in drug metabolism, detoxification of environmental toxins, and biotransformation of endogenous transmitters, has been implicated as a risk factor in neurodegenerative diseases. It has been observed to play a role in the development of both Parkinson’s disease and Alzheimer’s disease, but its role in ALS has not been established.

Using whole-genome sequencing data from more than 6000 individuals in the multinational Project MinE consortium, we examined whether *CYP2D6* variants and genotype-inferred metaboliser phenotypes influence ALS risk and survival. We used both multivariable logistic regression and multivariable Cox proportional hazards regression for the analyses, controlling for important clinical covariates.

Reduced CYP2D6 activity, driven by the common loss-of-function *CYP2D6*4* and other *-alleles causing either decrease or loss of the enzyme function, was associated with increased risk of ALS. Although *CYP2D6* variation had no overall effect on survival, in patients receiving Riluzole we observed a protective association, with poor metabolisers showing the greatest survival advantage compared with normal metabolisers.

These findings suggest that variation in *CYP2D6* contributes to ALS susceptibility and can modify treatment response. Incorporating *CYP2D6* genetic profiling into ALS clinical trials could reduce pharmacokinetic variability and improve detection of therapeutic effects. More broadly, this work provides a rationale for integrating pharmacogenomics into ALS research and care as a step towards precision medicine in neurodegeneration.

## Introduction

Amyotrophic lateral sclerosis (ALS) is a fatal neurodegenerative disorder characterised by progressive loss of upper and lower motor neurons.^1^ Despite decades of research, the causes remain only partly understood. Established risk factors include age, male sex, family history, and environmental exposures.^2–5^ Genetic contributions are clear, with hexanucleotide repeat expansion of *C9orf72* and mutations in *SOD1, TARDBP,* and *FUS* among the most common risk variants,^3^ yet known genes account for only up to 20% of apparently sporadic ALS.^6–8^ Moreover, genetic influences on disease risk and progression do not always overlap,^9,10^ underscoring the complexity of ALS pathogenesis.

Cytochrome P450 2D6 (CYP2D6) is one of the most important drug-metabolising enzymes, responsible for metabolising around a fifth of all commonly prescribed drugs.^11^ The gene encoding for CYP2D6 is highly polymorphic, with more than 185 recognised *\*(star*)-alleles to date,^12^ giving rise to phenotypes ranging from poor to ultrarapid metabolisers.^13,14^ CYP2D6 ultrarapid metabolisers metabolise substrates faster, while poor metabolisers have no enzyme activity. The most common loss-of-function variant among Europeans is a G/A transition at the intron 3- exon 4 junction *(CYP2D6*4)*, present in about 18-20% of individuals.^14,15^ This genetic variation in *CYP2D6* is also clinically relevant: altered enzyme activity contributes to drug toxicity, treatment failure, and variable therapeutic response.

Beyond drug metabolism, CYP2D6 plays a key role in detoxifying environmental toxins implicated in neurodegeneration. Reduced CYP2D6 activity has been consistently linked to increased risk of Parkinson’s disease,^16–19^ probably through impaired clearance of MPTP, pesticides, and other related neurotoxins.^20^ Poor CYP2D6 metabolism has also been proposed as a risk modifier in Alzheimer’s disease.^21^

In ALS, evidence of CYP2D6 involvement is less clear. Early small studies suggested an enrichment of *CYP2D6*4* among patients,^22^ but subsequent larger studies failed to replicate these findings.^23^ Given the mechanistic overlap with Parkinson’s disease and the role of environmental toxins in ALS risk, a more comprehensive analysis of *CYP2D6* genetic variation in ALS is warranted. We therefore investigated whether *CYP2D6* variants and genotype-inferred metaboliser phenotypes influence ALS risk or survival, using whole-genome sequencing data from the multinational Project MinE ALS Sequencing Consortium.

## Materials and methods

### Data description

We analysed whole-genome sequencing samples from 4140 ALS cases and 1812 controls from six countries from the Project MinE ALS Sequencing Consortium^24^ for the risk association analysis, 4459 cases from eight countries for survival analyses, and 1241 cases from four countries for the subset analysis where Riluzole use at time of blood draw was known. People were diagnosed with ALS according to the revised El Escorial Criteria^25^ at motor neuron disease clinics by neurologists specialized in motor neuron disease. Participants were from various European ancestries. We included people with ALS regardless of family history of ALS or dementias. All participants gave their informed consent.

### Genotyping *CYP2D6*

We used Cyrius(v1.1.1)^26^ to genotype the *CYP2D6* gene. The Cyrius tool has been designed to genotype the *CYP2D6* gene and can overcome the difficulties associated with genotyping this specific locus, achieving the highest accuracy compared to other genotyping tools with the capacity to genotype the *CYP2D6* gene.^26^

### Calculating *CYP2D6*\*4 count

The Cyrius tool is very effective for typing both structural variants and small or single nucleotide variants at the *CYP2D6* locus. There are many *CYP2D6* structural variants, such as a deletions, insertions or an additional copies of the gene. Therefore, the *CYP2D6*\*4 count must consider if the copies of *4 are situated on the same or two different alleles. Here we have calculated haplotype *CYP2D6*\*4 count naïve and total diplotype *CYP2D6*\*4 alleles. We used the naïve *4 allele counts for our analyses for two reasons. First, as the *4 allele represents a loss of function phenotype and will therefore not increase the metabolising activity of the enzyme, multiple copies of *4 on one allele make no functional difference. Secondly, the numbers of people with more than two copies of *CYP2D6*\*4 were comparatively too small across our whole dataset to capture any effect with sufficient power.

### Inferring the gene activity and phenotype

The Clinical Pharmacogenetics Implementation Consortium has created and maintains a database of the associations of each of more than 185 recognised *CYP2D6* *-alleles to the enzyme activity score.^27^ These activity scores are assigned to haplotypes and summed to calculate diplotype activity score. This diplotype activity score can in turn be used to infer the metaboliser phenotype: poor metabolisers have an activity score of 0.0, intermediate metabolisers a score between 0.25-1.0, normal or extensive metabolisers a score between 1.25- 2.25, and ultrarapid metabolisers a score above 2.25.

We have written an algorithm to automatically calculate the *CYP2D6* genotype-based activity score to infer the metaboliser phenotype. This algorithm is written in Python (version 3.8.0) and can be used to match the output files from the Illumina Cyrius tool directly to the Clinical Pharmacogenetics Implementation Consortium assigned activity scores and the generalised metaboliser phenotypes. The algorithm is publicly available on <u>github.com/johannakristiina/CYP2D6_phenotype_calculator</u>.

### Data cleaning

#### Risk analyses

We only studied samples from blood, excluding sequences from other tissues (*n* = 297). Where sample type was not specified, we assumed blood. We excluded samples with missing phenotype information (*n* = 371), and any samples which had discrepancies in reported and genetic sex (*n* = 36). We also removed samples with missing age at blood draw (*n* = 1201). *CYP2D6* genotyping was successful in all participants, however as we were interested in the genotypes with known effects, we removed all samples which included genotypes of unknown effects (*n* = 374). Finally, we removed cohorts which had either only cases or only controls data available (*n* = 159).

#### Survival analyses

We discarded samples not sequenced from blood (*n* = 297), assuming all samples with unspecified tissue type to be from blood. Next, we excluded all controls, retaining only ALS cases. We then removed any samples which had discrepancies between reported and genetic sex (*n* = 22). We also removed samples with missing age at onset (*n* = 93), and any samples with missing survival information (*n* = 92). We removed samples which included *CYP2D6* genotypes of unknown effects (*n* = 295). We then excluded samples where site of onset was set as FTD (*n* = 1) and removed samples where *C9orf72* expansion status information was missing (*n* = 142). Additionally, we performed analyses in a subset where Riluzole use at blood draw was known, removing samples where this information was unavailable (*n* = 2941). As we only had information if an individual had been on Riluzole at time of blood draw, and not if they had taken Riluzole at a later time, we only included those where Riluzole use at blood draw was confirmed (*n* = 1241).

### Population analyses

We calculated the frequency of *CYP2D6*\*4 allele and genotype inferred metaboliser phenotypes across our cohorts and within each cohort. We did this to assess any discrepancies in our cohort and previously reported respective frequencies in European cohorts.

### Statistical methods and survival analyses

We used the *CYP2D6*4* count and genotype-inferred metaboliser phenotypes along with relevant clinical information and survival times to analyse the association of *CYP2D6* variation with ALS risk and survival. Survival time was calculated from self-reported age at onset to age at death or last follow up. We used multivariable logistic regression and multivariable Cox Proportional Hazards models. In the multivariable logistic regression for ALS risk, we added covariates to control for age at onset, sex, sequencing technology, country, and the first 20 principal components for ancestry. We additionally controlled for site of onset and *C9orf72* expansion status in the Cox model as these have been shown to have strong effects on survival in ALS. Both multivariable logistic regression and multivariable Cox models were run first across cohorts, then per cohort, and meta-analysed across cohorts. False discovery rate (*P* < 0.05) correction was applied to the across all cohorts analyses, per cohort analyses, and across-cohort meta-analyses.

### Reproducibility

Data was cleaned and prepared for analysis using R (version 4.4.3). Logistic regression was run using the generalised linear model function from R stats (version 4.4.3). Survival analyses were run using integrated R packages survival (version 3.8.3) and survminer (version 0.5.0). Meta-analysis was run using the R metafor library (version 4.8.0).

## Results

### Study population overview and *CYP2D6*\*4 frequency among people with ALS and controls

In our analysis subset of Project MinE, only 84 people with ALS had a family member with the disease, while most (*n* = 4684) had the sporadic form. In our dataset, the frequencies of *CYP2D6*\*4 were consistent with previous literature with a mean frequency around 0.18-0.20 in Europeans.^14^ We observed similar *CYP2D6*\*4 frequencies both in people with ALS and healthy controls across cohorts (*t* = -1.9, *P* = 0.052). Overview of the *CYP2D6*\*4 frequencies and details of other relevant population statistics are presented in Table 1.

**Table 1.** Study population overview with *CYP2D6*4* allele frequencies and CYP2D6 metaboliser phenotype percentages.

|  | *n* Samples |  | Age (years)<sup>a</sup> |  | %Male |  | CYP2D6*4 freq. |  | %PM |  | %IM |  | %NM |  | %UM |  |
| --- | --- | --- | --- | --- | --- | --- | --- | --- | --- | --- | --- | --- | --- | --- | --- | --- |
|  | ALS | Controls | ALS | Controls | ALS | Controls | ALS | Controls | ALS | Controls | ALS | Controls | ALS | Controls | ALS | Controls |
| **Across cohorts** | 4929 | 1812 | 63.1 | 61.9 | 60.1 | 52.6 | 0.20 | 0.18 | 6.9 | 6.2 | 41.5 | 39.6 | 49.4 | 52.5 | 2.2 | 1.7 |
| Netherlands | 1687 | 964 | 64.6 | 63.7 | 59.0 | 58.9 | 0.19 | 0.17 | 6.2 | 5.7 | 45.1 | 40.8 | 46.9 | 51.9 | 1.8 | 1.7 |
| UK | 1219 | 380 | 62.4 | 60.1 | 61.9 | 38.7 | 0.20 | 0.18 | 7.1 | 6.6 | 40.9 | 37.9 | 50.5 | 53.4 | 1.6 | 2.1 |
| Ireland | 452 | 228 | 64.6 | 64.2 | 60.2 | 59.2 | 0.19 | 0.22 | 6 | 7.9 | 40.5 | 41.2 | 52.7 | 50.4 | 0.9 | 0.4 |
| US | 372 | 78 | 59.8 | 56.1 | 61.8 | 37.2 | 0.22 | 0.21 | 10.8 | 5.1 | 36.8 | 51.3 | 51.1 | 43.6 | 1.3 | 0 |
| Spain | 300 | 124 | 57.7 | 53.1 | 58.7 | 46.0 | 0.21 | 0.17 | 6.3 | 7.3 | 40.3 | 29.8 | 45.3 | 58.9 | 8 | 4 |
| France | 243 | 38 | 65.6 | 63.2 | 58.9 | 47.4 | 0.17 | 0.12 | 6.2 | 5.3 | 35 | 23.7 | 55.1 | 68.4 | 3.7 | 2.6 |
| Belgium | 494 | NA | NA | NA | 60.3 | NA | 0.21 | NA | 8.9 | NA | 42.5 | NA | 46 | NA | 2.6 | NA |
| Sweden | 162 | NA | NA | NA | 56.2 | NA | 0.13 | NA | 3.7 | NA | 30.2 | NA | 63 | NA | 3.1 | NA |

IM – intermediate metaboliser, NM – normal metaboliser, PM – poor metaboliser, UM – ultrarapid metaboliser.
<sup>a</sup>Age at blood draw.

**Table 2.** Results from evaluating CYP2D6*4 genotype on ALS risk.

|  | *n* ALS | *n* Controls | Recessive |  |  | Dominant |  |  |
| --- | --- | --- | --- | --- | --- | --- | --- | --- |
|  |  |  | *n* | OR (95%CI) | Adj. *P* | *n* | OR (95%CI) | Adj. *P* |
| **Logistic regression** | 4140 | 1812 | 5952 | 1.25 (0.86-1.77) | 0.26 | 5952 | 1.16 (1.03-1.30) | 0.03* |
| **Meta-analysis** | 4140 | 1812 | 5952 | 1.16 (0.80-1.68) | 0.43 | 5952 | 1.15 (1.02-1.30) | 0.04* |
| Netherlands | 1668 | 964 | 2632 | 1.26 (0.66-2.39) | 0.65 | 2632 | 1.21 (1.02-1.43) | 0.35 |
| UK | 1211 | 380 | 1591 | 1.58 (0.78-3.22) | 0.65 | 1591 | 1.18 (0.92-1.53) | 0.65 |
| Ireland | 395 | 228 | 623 | 0.68 (0.30-1.53) | 0.65 | 623 | 0.85 (0.60-1.21) | 0.65 |
| Spain | 298 | 124 | 422 | 1.33 (0.39-4.49) | 0.76 | 422 | 1.20 (0.74-1.93) | 0.65 |
| US | 329 | 78 | 407 | 1.99 (0.40-9.90) | 0.65 | 407 | 1.11 (0.63-1.96) | 0.76 |
| France | 239 | 38 | 277 | 1.43 (0.15-13.45) | 0.76 | 277 | 1.71 (0.71-4.12) | 0.65 |

Across cohorts analyses are noted in bold.
\*FDR *P* < 0.05

### Investigating *CYP2D6*\*4 genotype on ALS risk

The multivariable logistic regression and the meta-analysis suggest an association of *CYP2D6*\*4 polymorphism with increased risk of developing ALS in the dominant model after multiple testing correction (*OR* = 1.16, *95% CI* = 1.03-1.30, *adj. P* = 0.03; meta-analysis *OR* = 1.15, *95% CI* = 1.02-1.30, *adj. P* = 0.04). The recessive model was not significant for the association with ALS risk in both the multivariable logistic regression or the meta-analysis (*OR* = 1.25, *95% CI* = 0.86-1.77, *adj. P* = 0.26; meta-analysis *OR* = 1.16, *95% CI* = 0.80-1.68, *adj. P* = 0.43). In the per cohort analyses, none of the evaluated models were significant after multiple testing correction (**Error! Reference source not found.**).

### CYP2D6 metaboliser phenotypes in the population

In the Project MinE dataset, we also evaluated the prevalence of different *CYP2D6* genotype inferred metaboliser phenotypes. These too follow the previously established percentages in European populations.^14^ We similarly observed that the frequencies of non-normal metaboliser phenotypes – poor metaboliser, intermediate metaboliser, and ultrarapid metaboliser – are similar between the control and ALS populations (*X*^2^ = 6.40, *df* = 3, *P* = 0.09) (Table 1).

### Investigating CYP2D6 metaboliser phenotype on ALS risk

Both the multivariable logistic regression and the meta-analysis suggest that any change from normal CYP2D6 metaboliser phenotype is associated with risk of developing ALS, as is observed in the model evaluating divergence from normal metaboliser phenotype (*OR* = 1.18, *95% CI* = 1.06-1.33, *adj. P* = 0.01; meta-analysis *OR* = 1.18, *95% CI* = 1.05-1.32, *adj. P* = 0.02). This seems to be driven by the effect of the decreased CYP2D6 metaboliser phenotypes, the poor metaboliser and intermediate metaboliser phenotypes, as observed in the model evaluating the combined decreased phenotypes against normal metabolisers (*OR* = 1.18, *95% CI* = 1.05- 1.33, *P* = 0.01, meta-analysis *OR* =1.18, *95% CI* = 1.05-1.33, *P* = 0.02). Details on all model results are presented in Table 3.

**Table 3.** Results from evaluating CYP2D6 metaboliser phenotype on ALS risk.

|  | **n Samples** |  | **Any non-NM** |  |  | **PM vs NM** |  |  | **IM vs NM** |  |  | **UM vs NM** |  |  | **PM+IM vs NM** |  |  |
| --- | --- | --- | --- | --- | --- | --- | --- | --- | --- | --- | --- | --- | --- | --- | --- | --- | --- |
|  | **ALS** | **Control** | **n** | **OR (95%CI)** | **Adj. p** | **n** | **OR (95%CI)** | **Adj. P** | **n** | **OR (95%CI)** | **Adj. P** | **n** | **OR (95%CI)** | **Adj. P** | **n** | **OR (95%CI)** | **Adj. P** |
| **Logistic regression** | 4140 | 1812 | 5952 | 1.18 (1.06 - 1.33) | 0.01* | 3381 | 1.21 (0.96 - 1.54) | 0.14 | 5439 | 1.18 (1.05 - 1.33) | 0.01* | 3108 | 1.25 (0.82 - 1.95) | 0.31 | 5832 | 1.18 (1.05 - 1.33) | 0.01* |
| **Meta-analysis** | 4140 | 1812 | 6657 | 1.18 (1.05 - 1.32) | 0.02* | 3381 | 1.20 (0.93 - 1.53) | 0.19 | 5439 | 1.18 (1.04 - 1.33) | 0.02* | 3108 | 1.08 (0.68 - 1.72) | 0.75 | 5832 | 1.18 (1.05 - 1.33) | 0.02* |
| Netherlands | 1668 | 964 | 2632 | 1.22 (1.04 - 1.43) | 0.19 | 1442 | 1.24 (0.87 - 1.77) | 0.45 | 2428 | 1.22 (1.03 - 1.45) | 0.19 | 1328 | 1.07 (0.56 - 2.04) | 0.87 | 2587 | 1.23 (1.04 - 1.45) | 0.19 |
| UK | 1211 | 380 | 1591 | 1.17 (0.92 - 1.49) | 0.42 | 924 | 1.30 (0.79 - 2.13) | 0.55 | 1453 | 1.19 (0.92 - 1.54) | 0.42 | 840 | 0.64 (0.26 - 1.56) | 0.58 | 1564 | 1.20 (0.94 - 1.53) | 0.42 |
| Ireland | 395 | 228 | 623 | 0.90 (0.64 - 1.27) | 0.64 | 364 | 0.73 (0.37 - 1.46) | 0.62 | 575 | 0.92 (0.64 - 1.32) | 0.71 | 326 | 2.83 (0.26 - 30.8) | 0.62 | 618 | 0.89 (0.63 - 1.25) | 0.64 |
| US | 298 | 124 | 422 | 1.59 (1.01 - 2.51) | 0.34 | 236 | 1.32 (0.53 - 3.32) | 0.64 | 365 | 1.63 (0.98 - 2.72) | 0.36 | 237 | 2.19 (0.75 - 6.41) | 0.42 | 393 | 1.52 (0.95 - 2.45) | 0.38 |
| Spain | 329 | 78 | 407 | 0.84 (0.48 - 1.46) | 0.64 | 241 | 1.45 (0.43 - 4.93) | 0.64 | 366 | 0.68 (0.38 - 1.21) | 0.42 | 208 | 3.45 (0.00 - Inf)<sup>a</sup> | 0.99 | 403 | 0.82 (0.47 - 1.43) | 0.64 |
| France | 239 | 38 | 277 | 1.80 (0.80 - 4.05) | 0.42 | 174 | 1.75 (0.31 - 9.83) | 0.64 | 252 | 2.20 (0.86 - 5.58) | 0.38 | 169 | 0.56 (0.05 - 6.09) | 0.71 | 267 | 2.03 (0.87 - 4.76) | 0.38 |

Across cohorts analyses are noted in bold. IM – intermediate metaboliser, NM – normal metaboliser, PM – poor metaboliser, UM – ultrarapid metaboliser.
<sup>a</sup>Not enough ultrarapid metabolisers in the Spanish cohort to measure any effect of this metaboliser phenotype on ALS risk.
\*FDR $P < 0.05$

### Investigating *CYP2D6*\*4 genotype on ALS survival

The pooled Cox model and meta-analysis suggest no effect of *CYP2D6*\*4 on survival in ALS. We observed no models to be significant on either across cohorts, per-cohort, or meta-analysis level (Table 4).

**Table 4.** Results from investigating *CYP2D6*4* genotype on ALS survival.

|  | Recessive |  |  | Dominant |  |  |
| --- | --- | --- | --- | --- | --- | --- |
|  | *n* | HR (95%CI) | Adj. *P* | *n* | HR (95%CI) | Adj. *P* |
| *All ALS* |  |  |  |  |  |  |
| **Multivariable Cox regression** | 4459 | 0.91 (0.75 - 1.11) | 0.36 | 4459 | 0.97 (0.91 - 1.04) | 0.36 |
| **Meta-analysis** | 4459 | 0.94 (0.77 - 1.14) | 0.52 | 4459 | 0.98 (0.91 - 1.05) | 0.52 |
| Netherlands | 1612 | 1.06 (0.71 - 1.58) | 0.99 | 1612 | 1.00 (0.90 - 1.11) | 0.99 |
| UK | 1035 | 1.00 (0.70 - 1.42) | 0.99 | 1035 | 1.04 (0.90 - 1.20) | 0.99 |
| Belgium | 480 | 0.81 (0.51 - 1.28) | 0.99 | 480 | 0.82 (0.66 - 1.01) | 0.8 |
| Ireland | 404 | 1.17 (0.61 - 2.24) | 0.99 | 404 | 0.98 (0.77 - 1.26) | 0.99 |
| US | 317 | 0.74 (0.35 - 1.58) | 0.99 | 317 | 1.06 (0.80 - 1.41) | 0.99 |
| Spain | 259 | 0.60 (0.21 - 1.73) | 0.99 | 259 | 0.97 (0.66 - 1.43) | 0.99 |
| France | 194 | 0.43 (0.08 - 2.23) | 0.99 | 194 | 0.88 (0.53 - 1.46) | 0.99 |
| Sweden | 158 | 0.79 (0.09 - 6.82) | 0.99 | 158 | 0.65 (0.39 - 1.08) | 0.8 |
| *On Riluzole* |  |  |  |  |  |  |
| **Multivariable Cox regression** | 1241 | 0.78 (0.54 - 1.12) | 0.17 | 1241 | 0.86 (0.75 - 0.98) | 0.05 |
| **Meta-analysis** | 1241 | 0.76 (0.52 - 1.10) | 0.15 | 1241 | 0.87 (0.76 - 1.01) | 0.12 |
| Netherlands | 287 | 0.54 (0.17 - 1.67) | 0.53 | 287 | 0.88 (0.67 - 1.15) | 0.53 |
| Belgium | 419 | 0.78 (0.48 - 1.27) | 0.53 | 419 | 0.81 (0.65 - 1.02) | 0.53 |
| Ireland | 317 | 1.04 (0.47 - 2.29) | 0.93 | 317 | 0.91 (0.68 - 1.21) | 0.68 |
| Spain | 218 | 0.35 (0.08 - 1.56) | 0.53 | 218 | 1.06 (0.68 - 1.66) | 0.90 |

Across cohorts analyses are noted in bold. Subgroup analyses are noted in italics.

### Investigating CYP2D6 metaboliser phenotype on ALS survival

In both, the across-cohorts Cox model and the meta-analysis, we did not observe any effect of CYP2D6 metaboliser phenotypes on survival in ALS. All model results are presented in Table 5.

**Table 5.** Results from exploring the CYP2D6 metaboliser phenotype effect on survival in ALS.

|  | Any non-NM |  |  | PM vs NM |  |  | IM vs NM |  |  | UM vs NM |  |  | PM & IM vs NM |  |  |
| --- | --- | --- | --- | --- | --- | --- | --- | --- | --- | --- | --- | --- | --- | --- | --- |
|  | *n* | HR (95%CI) | Adj. *P* | *n* | HR (95%CI) | Adj. *P* | *n* | HR (95%CI) | Adj. *P* | *n* | HR (95%CI) | Adj. *P* | *n* | HR (95%CI) | Adj. *P* |
| *All ALS* |  |  |  |  |  |  |  |  |  |  |  |  |  |  |  |
| **Multivariable Cox regression** | 4459 | 0.97 (0.91 - 1.04) | 0.45 | 2504 | 0.95 (0.83 - 1.09) | 0.45 | 4063 | 0.97 (0.90 - 1.03) | 0.45 | 2304 | 1.17 (0.93 - 1.47) | 0.45 | 4361 | 0.97 (0.90 - 1.03) | 0.45 |
| **Meta-analysis** | 4459 | 0.98 (0.89 - 1.07) | 0.76 | 2504 | 0.89 (0.72 - 1.11) | 0.76 | 4063 | 0.99 (0.91 - 1.07) | 0.78 | 2304 | 1.23 (0.96 - 1.58) | 0.53 | 4361 | 0.97 (0.89 - 1.07) | 0.76 |
| Netherlands | 1612 | 0.97 (0.88 - 1.08) | 0.86 | 854 | 1.04 (0.84 - 1.30) | 0.89 | 1485 | 0.96 (0.86 - 1.06) | 0.75 | 785 | 1.20 (0.82 - 1.78) | 0.67 | 1583 | 0.97 (0.87 - 1.07) | 0.78 |
| UK | 1035 | 1.14 (1.00 - 1.31) | 0.51 | 600 | 1.22 (0.92 - 1.61) | 0.51 | 952 | 1.11 (0.96 - 1.28) | 0.51 | 545 | 1.59 (0.90 - 2.83) | 0.51 | 1021 | 1.13 (0.98 - 1.29) | 0.51 |
| Belgium | 480 | 0.87 (0.72 - 1.07) | 0.51 | 264 | 0.77 (0.52 - 1.13) | 0.51 | 424 | 0.86 (0.70 - 1.07) | 0.51 | 234 | 1.68 (0.87 - 3.25) | 0.51 | 467 | 0.84 (0.69 - 1.03) | 0.51 |
| Ireland | 404 | 0.94 (0.75 - 1.19) | 0.87 | 240 | 0.76 (0.45 - 1.27) | 0.67 | 376 | 0.98 (0.77 - 1.25) | 0.93 | 220 | 0.39 (0.11 - 1.39) | 0.51 | 400 | 0.96 (0.76 - 1.21) | 0.89 |
| US | 317 | 0.97 (0.73 - 1.30) | 0.93 | 189 | 0.83 (0.49 - 1.43) | 0.78 | 278 | 1.05 (0.76 - 1.45) | 0.89 | 160 | 1.74 (0.56 - 5.37) | 0.67 | 312 | 0.97 (0.72 - 1.30) | 0.93 |
| Spain | 259 | 1.08 (0.74 - 1.56) | 0.89 | 135 | 0.24 (0.08 - 0.74) | 0.40 | 222 | 1.18 (0.78 - 1.76) | 0.75 | 142 | 1.01 (0.52 - 1.97) | 0.98 | 237 | 1.03 (0.69 - 1.53) | 0.93 |
| France | 194 | 0.70 (0.42 - 1.18) | 0.51 | 117 | 0.97 (0.33 - 2.88) | 0.98 | 178 | 0.90 (0.51 - 1.58) | 0.89 | 113 | 0.06 (0.01 - 0.70) | 0.40 | 188 | 0.84 (0.50 - 1.41) | 0.78 |
| Sweden | 158 | 0.78 (0.50 - 1.20) | 0.63 | 105 | 0.54 (0.16 - 1.78) | 0.67 | 148 | 0.80 (0.50 - 1.27) | 0.67 | 105 | 1.27 (0.31 - 5.26) | 0.89 | 153 | 0.77 (0.49 - 1.20) | 0.63 |
| *On Riluzole* |  |  |  |  |  |  |  |  |  |  |  |  |  |  |  |
| **Multivariable Cox regression** | 1241 | 0.86 (0.76 - 0.98) | 0.03* | 681 | 0.67 (0.51 - 0.88) | 0.02* | 1117 | 0.88 (0.77 - 1.00) | 0.06 | 635 | 1.16 (0.78 - 1.72) | 0.46 | 1202 | 0.85 (0.75 - 0.96) | 0.03* |
| **Meta-analysis** | 1241 | 0.87 (0.76 - 1.00) | 0.08 | 681 | 0.65 (0.48 - 0.87) | 0.02* | 1117 | 0.90 (0.78 - 1.03) | 0.16 | 635 | 1.15 (0.42 - 3.16) | 0.79 | 1202 | 0.85 (0.75 - 0.98) | 0.06 |
| Netherlands | 287 | 0.80 (0.62 - 1.03) | 0.32 | 150 | 0.63 (0.34 - 1.19) | 0.32 | 269 | 0.82 (0.63 - 1.07) | 0.32 | 138 | 5.51 (1.37 - 22.18) | 0.20 | 284 | 0.78 (0.61 - 1.01) | 0.32 |
| Belgium | 419 | 0.89 (0.72-1.10) | 0.39 | 237 | 0.75 (0.49 - 1.14) | 0.32 | 369 | 0.90 (0.71 - 1.12) | 0.43 | 209 | 1.27 (0.60 - 2.69) | 0.59 | 408 | 0.87 (0.70 - 1.08) | 0.32 |
| Ireland | 317 | 0.83 (0.63 - 1.09) | 0.32 | 182 | 0.64 (0.34 - 1.19) | 0.32 | 294 | 0.86 (0.65 - 1.13) | 0.39 | 167 | 0.28 (0.07 - 1.14) | 0.32 | 313 | 0.84 (0.64 - 1.10) | 0.32 |
| Spain | 218 | 1.18 (0.77 - 1.82) | 0.52 | 112 | 0.17 (0.05 - 0.64) | 0.20 | 185 | 1.39 (0.86 - 2.24) | 0.32 | 121 | 0.92 (0.45 - 1.89) | 0.82 | 197 | 1.13 (0.71 - 1.81) | 0.63 |

Across cohorts analyses are noted in bold. Subgroup analyses are noted in italics. IM – intermediate metaboliser, NM – normal metaboliser, PM – poor metaboliser, UM – ultrarapid metaboliser.
\*FDR *P* < 0.05

### Investigating *CYP2D6*4* genotype on ALS survival within the group where Riluzole use is confirmed

We also performed a sub-group analysis for a group where Riluzole use at blood draw was known. In this subgroup analysis we did not observe any significant effect of *CYP2D6*\*4 genotype on ALS survival after multiple testing correction (Table 4).

### Investigating CYP2D6 metaboliser phenotype effect on ALS survival within the group where Riluzole use is confirmed

Inversely to the risk association, in the Riluzole subgroup analysis we observed poor *CYP2D6* metabolisers to have a small protective effect against earlier mortality in ALS. We observed significant effects in the across-cohorts Cox model evaluating any divergence from normal metaboliser phenotype (*HR* = 0.86, *95% CI* = 0.76-0.98, *adj. P* = 0.03). This effect was however not significant in the meta-analysis after multiple testing correction (*HR* = 0.87, *95% CI* = 0.76- 1.00, *adj. P* = 0.08). Similarly, in the model evaluating decreased activity metaboliser phenotypes, poor metabolisers and intermediate metabolisers, against normal metabolisers, we observed a significant effect in the across-cohorts Cox model (*HR* = 0.85, *95% CI* = 0.75-0.96, *adj. P* = 0.03), but not in the meta-analysis after multiple testing correction (*HR* = 0.85, *95% CI* = 0.75-0.98, *adj. P* = 0.06). However, in the model evaluating only the CYP2D6 poor metaboliser phenotype against the normal metaboliser phenotype, we observed evidence of a protective association in both the across-cohorts Cox analysis and the meta-analysis (*HR* = 0.67, *95% CI* = 0.51–0.88, *adj. P* = 0.02; meta-analysis *HR* = 0.65, *95% CI* = 0.48–0.87, *adj. P* = 0.02) (Table 5).

## Discussion

In this large, whole-genome sequencing study of more than 6000 individuals, we found that reduced *CYP2D6* activity was associated with increased risk of ALS. Although *CYP2D6* variation had no effect on survival in the overall cohort, a protective association was observed among patients treated with Riluzole, with the strongest effect in poor metabolisers. These findings suggest that pharmacogenetic variation at the *CYP2D6* locus contributes to ALS susceptibility and may modify treatment response.

Our results are consistent with evidence from other neurodegenerative disorders. In Parkinson’s disease, reduced *CYP2D6* activity has been linked to increased risk,^16–19^ possibly through impaired detoxification of neurotoxins,^20^ while associations in Alzheimer’s disease have been less consistent.^21^ The present analysis is the largest to assess *CYP2D6* in ALS, and the first to do so using whole-genome sequencing across multiple cohorts. This approach enabled robust characterisation of both structural and sequence variation, providing greater resolution than previous studies at this locus.

The survival advantage observed in Riluzole-treated poor metabolisers is clinically notable. Riluzole is the most widely used therapy for ALS, but its modest effect size has limited clinical utility. Impaired metabolism may prolong drug exposure, thereby amplifying benefit in specific patient subgroups. If validated, *CYP2D6* profiling could identify individuals more likely to respond to Riluzole, inform treatment decisions, and optimise trial design by reducing pharmacokinetic variability.

Beyond Riluzole, many experimental ALS therapies are metabolised through the cytochrome P450 system. Pharmacogenetic heterogeneity may contribute to the inconsistent efficacy signals seen in recent trials. Stratifying participants by *CYP2D6* or other pharmacogenes could improve power to detect treatment effects, reduce trial failure rates, and accelerate therapeutic development. Such strategies are now routine in oncology but remain underused in neurodegenerative disease. Our findings provide a rationale for integrating pharmacogenomics into ALS research and clinical practice as the field moves towards precision medicine.

These results also raise mechanistic questions. *CYP2D6* is involved in detoxifying pesticides and other xenobiotics that have been implicated in ALS.^5,28^ Reduced enzymatic activity could increase vulnerability to neurotoxic exposures, thereby contributing to risk. Although we lacked exposure data to test this directly, gene-environment interactions remain a plausible pathway and warrant further study.

Our analysis has limitations. First, data were restricted to individuals of European ancestry, limiting generalisability. Inclusion of more diverse populations is essential both to validate these findings and to ensure pharmacogenetic approaches are globally relevant. Second, although whole-genome sequencing and the Cyrius algorithm allow comprehensive genotyping, PCR remains more accurate for certain alleles, and some misclassification cannot be excluded. Third, treatment information was limited to Riluzole use at the time of sampling, without longitudinal data on dose or adherence. Finally, survival analyses were observational and residual confounding remains possible.

Nevertheless, the scale of this study, the use of whole-genome sequencing, and the consistency of associations across multiple analyses strengthen confidence in the findings. Replication in independent and multi-ethnic cohorts, ideally with pharmacokinetic data, will be critical. If confirmed, *CYP2D6* profiling could be incorporated into future ALS clinical trials and clinical care to guide treatment decisions.

In conclusion, *CYP2D6* variation appears to contribute both to ALS risk and to modification of therapeutic response. As new therapies emerge, pharmacogenetic stratification could play a central role in maximising efficacy and reducing trial attrition. Incorporating pharmacogenomics into ALS research offers a tangible step towards precision medicine in a field where effective treatments remain urgently needed.

## Data availability

The data sets used that support the findings in this study are available from The Project MinE consortium public repository. To gain access to the data, an account request must be made to. Data access will require the completion of a data access request. Further information about data access can be found at https://www.projectmine.com/datasharing/.

## Acknowledgements

AAC is an NIHR Senior Investigator (NIHR202421) and a Visiting Professor at the Perron Institute for Neurological and Translational Science, Australia.

## Project MinE Consortium

Alfredo Iacoangeli, Ramona A. J. Zwamborn, Wouter van Rheenen, Maarten Kooyman, Ross Byrne, Kristel R. Van Eijk, Kevin Kenna, Joke J.F.A. van Vugt, Johnathan Cooper-Knock, Brendan Kenna, Atay Vural, Simon Topp, Yolanda Campos, Markus Weber, Bradley Smith, Richard Dobson, Michael A. van Es, Peter Andersen, Patrick Vourc’h, Philippe Corcia, Philippe Couratier, Mamede de Carvalho, Marc Gotkine, Vivian Drory, Yossef Lerner, Monica P. Panades, Teresa Salas, Jesus S. Mora, Monica Povedano, Adriano Chio, Vincenzo Silani, Nicola Ticozzi,, Fleur Garton, Allan McRae, Pamela J. Shaw, Karen E. Morrison, John E. Landers, Jonathan D. Glass, Clifton L. Dalgard, Christopher E Shaw, Nazli Basak, Orla Hardiman, Philip Van Damme, Russell L. McLaughlin, Leonard H. van den Berg, Jan H. Veldink, Ammar Al-Chalabi, and Ahmad Al Khleifat.

The full list of all Project MinE data freeze 2 collaborators with affiliations can be viewed in supplementary file Appendix1.

