## Appendix 1 for "CYP2D6 variants in amyotrophic lateral sclerosis: an association study of risk and survival"

| **country** | **name** | | | |
|---|---|---|---|---|
| Belgium | Philip | | van | Damme |
| France | Philippe | | | Corcia |
| France | Philippe | | | Couratier |
| France | Patrick | | | Vourc'h |
| Ireland | Orla | | | Hardiman |
| Ireland | Russell | | | McLaughin |
| Israel | Marc | | | Gotkine |
| Israel | Yossef | | | Lerner |
| Israel | Vivian | | | Drory |
| Italy | Nicola | | | Ticozzi |
| Italy | Vincenzo | | | Silani |
| NL | Jan | H. | | Veldink |
| NL | Leonard | H. | van den | Berg |
| Portugal | Mamede | | de | Carvalho |
| Spain | Teresa | | | Salas |
| Spain | Jesus | S. | | Mora Pardina |
| Spain | Monica | | | Povedano |
| Sweden | Peter | | | Andersen |
| Switzerland | Markus | | | Weber |
| Turkey | Nazli | A. | | Başak |
| UK | Ammar | | | Al-Chalabi |
| UK | Chris | | | Shaw |
| UK | Pamela | J. | | Shaw |
| UK | Karen | E. | | Morrison |
| USA | John | E. | | Landers |
| USA | Jonathan | D. | | Glass |
| USA | Clifton | L. | | Dalgard |
| UK | Jonathan | | | Cooper-Knock |
| UK | Ahmad | | | Al Khleifat |
| UK | Alfredo | | | Iacoangeli |
| NL | Wouter | | | van Rheenen |
| NL | Ramona | | | Zwamborn |
| NL | Joke | | | van Vugt |
| NL | Maarten | | | Kooyman |
| NL | Kevin | | | Kenna |
| NL | Michael | | | van Es |
| Italy | Adriano | | | Chio |

| email | DF2 |
| --- | --- |
| | Y |
| | Y |
| | Y |
| | Y |
| | Y |
| | Y |
| | Y |
| | Y |
| | Y |
| | *Y* |
| | Y |
| | Y |
| | Y |
| | Y |
| | Y |
| | Y |
| | Y |
| | Y |
| | Y |
| | Y |
| | Y |
| | Y |
| | Y |
| | Y |
| | Y |
| | Y |
| | Y |
| | |
| | |
| | |
| | |
| | |
| | |
| | |
| | |
| | |
| | |

**affiliations (may change in time!)**

KU Leuven - University of Leuven, Department of Neurosciences
Centre SLA, CHRU de Tours, Tours, France; UMR 1253, iBrain, Université de Tours, Inserm, Tours, France.
Centre SLA CHU Dupuytren Limoges France.
Service de Biochimie et Biologie moléculaire, CHU de Tours, Tours, France
Academic Unit of Neurology, Trinity College Dublin, Trinity Biomedical Sciences Institute, Dublin, Republic of Ir
Complex Trait Genomics Laboratory, Smurfit Institute of Genetics, Trinity College Dublin, Dublin, Republic of Ir
Department of Neurology, Hadassah Medical Organization and Faculty of Medicine, Hebrew University of Jeru
Department of Neurology, Hadassah Medical Organization and Faculty of Medicine, Hebrew University of Jeru
Department of Neurology Tel-Aviv Sourasky Medical Centre , Israel.
Department of Neurology and Laboratory of Neuroscience, IRCCS Istituto Auxologico Italiano, Milano, Italy.
Department of Neurology and Laboratory of Neuroscience, IRCCS Istituto Auxologico Italiano, Milano, Italy.
Department of Neurology, UMC Utrecht Brain Center, University Medical Center Utrecht, Utrecht University, U
Department of Neurology, UMC Utrecht Brain Center, University Medical Center Utrecht, Utrecht University, U
Instituto de Fisiologia, Instituto de Medicina Molecular,Faculdade de Medicina, Universidade de Lisboa, Lisbor
ALS Unit, Hospital Universitario La Paz-Carlos III, Madrid, Spain
ALS Unit, Hospital San Rafael, Madrid, Spain.
la Unitat Funcional de Motoneurona, Cap de Secció de Neurofisiologia, Servei de Neurologia, Hospital Universi
Department of Clinical Science, Neurosciences, Umeå University, Sweden.
Neuromuscular Diseases Unit/ALS Clinic, Kantonsspital St. Gallen, 9007, St. Gallen, Switzerland.
Koç University, School of Medicine, Molecular Biology and Genetics- KUTTAM, Suna and Inan Kıraç Foundation
Maurice Wohl Clinical Neuroscience Institute, King's College London, Department of Basic and Clinical Neurosc
Maurice Wohl Clinical Neuroscience Institute, King's College London, Department of Basic and Clinical Neurosc
Sheffield Institute for Translational Neuroscience (SITraN), University of Sheffield, Sheffield, UK.
School of Medicine, Dentistry and Biomedical Sciences, Queen's University Belfast, UK.
Department of Neurology, University of Massachusetts Medical School, Worcester, MA, USA.
Department Neurology, Emory University School of Medicine, Atlanta, GA, USA.
The American Genome Center, Uniformed Services University - "America's Medical School", Bethesda, MD, US
Sheffield Institute for Translational Neuroscience (SITraN), University of Sheffield, Sheffield, UK.
Maurice Wohl Clinical Neuroscience Institute, King's College London, Department of Basic and Clinical Neurosc
Maurice Wohl Clinical Neuroscience Institute, King's College London, Department of Basic and Clinical Neurosc
Department of Neurology, UMC Utrecht Brain Center, University Medical Center Utrecht, Utrecht University, U
Department of Neurology, UMC Utrecht Brain Center, University Medical Center Utrecht, Utrecht University, U
Department of Neurology, UMC Utrecht Brain Center, University Medical Center Utrecht, Utrecht University, U
Department of Neurology, UMC Utrecht Brain Center, University Medical Center Utrecht, Utrecht University, U
Department of Neurology, UMC Utrecht Brain Center, University Medical Center Utrecht, Utrecht University, U
Department of Neurology, UMC Utrecht Brain Center, University Medical Center Utrecht, Utrecht University, U
Universita degli Studi dei Torino, Italy

Federation des Centres SLA Tours and Limoges, LITORALS, Tours, France.
Federation des Centres SLA Tours and Limoges, LITORALS, Tours, France.
UMR 1253, Université de Tours, Inserm, 37044 Tours, France
Department of Neurology, Beaumont Hospital, Dublin, Republic of Ireland.
NA
NA
salem, Israel
NA
Department of Pathophysiology and Tranplantation, 'Dino Ferrari' Center, Università degli Studi di Milan
Department of Pathophysiology and Tranplantation, 'Dino Ferrari' Center, Università degli Studi di Milan
NA
NA
Department of Neurosciences, Hospital de Santa Maria-CHLN, Lisbon, Portugal.

tario de Bellvitge-IDIBELL
NA
NA
NA
Department of Neurology, King's College Hospital, London SE5 9RS, UK
NA
NA
NA
NA
Emory ALS Center, Emory University School of Medicine, Atlanta, GA, USA.
SA

cience, London, UK.
cience, London, UK.
Utrecht, The Netherlands.
Utrecht, The Netherlands.
Utrecht, The Netherlands.
Utrecht, The Netherlands.
Utrecht, The Netherlands.
Utrecht, The Netherlands.

no, Milano, Italy.
no, Milano, Italy.

| country | name | Column1 | Column2 | Column3 | email |
|---|---|---|---|---|---|
| Belgium | Philip | | van | Damme | |
| France | Philippe | | | Corcia | |
| France | Philippe | | | Couratier | |
| France | Patrick | | | Vourc'h | |
| Ireland | Orla | | | Hardiman | |
| Ireland | Russell | | | McLaughin | |
| Israel | Marc | | | Gotkine | |
| Israel | Yossef | | | Lerner | |
| Israel | Shovman | | | Yehuda | |
| Israel | Vivian | | | Drory | |
| Italy | Nicola | | | Ticozzi | |
| Italy | Vincenzo | | | Silani | |
| NL | Jan | H. | | Veldink | |
| NL | Leonard | H. | van den | Berg | |
| Portugal | Mamede | | de | Carvalho | |
| Spain | Teresa | | | Salas | |
| Spain | Jesus | S. | | Mora Pardina | |
| Spain | Monica | | | Povedano | |
| Sweden | Peter | | | Andersen | |
| Switzerland | Markus | | | Weber | |
| Turkey | Nazli | A. | | Başak | |
| UK | Ammar | | | Al-Chalabi | |
| UK | Chris | | | Shaw | |
| UK | Pamela | J. | | Shaw | |
| UK | Karen | E. | | Morrison | |
| USA | John | E. | | Landers | |
| USA | Jonathan | D. | | Glass | |
| USA | Clifton | L. | | Dalgard | |

| DF2 |
| --- |
| Y |
| Y |
| Y |
| Y |
| Y |
| Y |
| Y |
| Y |
| Y |
| Y |
| Y |
| Y |
| Y |
| Y |
| Y |
| Y |
| Y |
| Y |
| Y |
| Y |
| Y |
| Y |
| Y |
| Y |
| Y |
| Y |
| Y |
| Y |
